# Experimental multi-center validation of a radiomics-based photonic quantum precision medicine architecture for lesion-level prediction of anti-PD-1 response in non-small cell lung cancer

**DOI:** 10.64898/2026.03.09.26347939

**Authors:** Stefano Olgiati, Federico Santona, Davide Meloni, Emanuele Barabino, Giovanni Rossi, Carlo Genova, Francesco Grossi, Nima Heidari

**Affiliations:** Università degli Studi di Ferrara, Department of Translational Medicine, Ferrara, IT; Quantum Spektral, Chivasso (TO), IT; Aarhus University, Aarhus, Central Jutland, DK; IRCCS Azienda Ospedaliera Metropolitana, Department of General Radiology, Genova, IT; IRCCS Azienda Ospedaliera Metropolitana, Department of Medical Oncology, Genova, IT; IRCCS Azienda Ospedaliera Metropolitana, Department of Academic Medical Oncology, Genova, IT; University of Medicine, Pharmacy, Science and Technology “G.E. Palade” of Târgu Mureş, RO

**Keywords:** immunotherapy, non-small cell lung cancer, radiomics, photonic quantum machine learning

## Abstract

**Background:** Previous research has shown that radiomics-based machine learning models are promising precision medicine tools for lesion-level predictions of Anti-PD-1 response in advanced non-small cell lung cancer (NSCLC) but their clinical implementation remains limited due to poor generalizability and uncertainty about which features capture true biological signals or merely reflect noise. We tested the performance and multi-center validity of a radiomics-based photonic quantum architecture trained on a feature space reduced using clinical knowledge and robust medical statistics.

**Methods:** This study included 125 patients with 164 advanced NSCLC single lesions from 3 different hospitals (Train, Test 1 and Test 2) treated with anti-PD1 monotherapy as first or second line. All patients underwent a baseline CT scan before the start of the treatment, the lesions were semi-automatically segmented and labeled as “progressive” if their diameter increased by more than 10% and as “non-progressive” if their diameter decreased by 10% or more over the following 6 months. From each CT scan we extracted 851 radiomic features with a METhodological RadiomICs Score (METRICS) of 86.1% (Excellent Quality Category (Table S1)) of which 183 were identified as reliable based on previous published clinical research. We then trained 1 classical and 3 photonic quantum machine learning models in Train Hospital and tested their performance on unseen external datasets in Test Hospitals 1 & 2. Aiming to explore quantum machine learning as a long-term technology for precision oncology, we utilized an ideal classical simulation of a photonic quantum architecture. This approach assumes perfectly functioning hardware, eliminating confounding physical noise to assess true theoretical performance. Crucially, by adapting the standardized MerLin template, we ensure our results are reproducible, fulfilling an essential requirement for evidence-based clinical research.

**Findings:** We found that only 2 features out of 851 were both reliable and robustly correlated to the target (*p* < 0.001). These 2 features were used to train the machine learning models. Across external validation datasets, the LEXGROUPING-6modes quantum architecture explicitly outperformed the classical MLP baseline in Test Hospital 1 (Average Precision 0.755 vs. 0.702) and matched its performance in Test Hospital 2 (0.670). Notably, all photonic quantum architectures successfully exceeded the chance level defined by progressor prevalence (0.622 and 0.462, respectively).

**Interpretation:** To our knowledge, this is the first study that tests the external validity of radiomics-based photonic quantum architectures utilizing an evidence-based, statistically significant reduced feature space. Crucially, demonstrating that a quantum architecture can outperform or match an optimized classical baseline represents a significant milestone. These findings validate the theoretical potential of quantum models to capture complex biological signals, supporting their future role as clinical decision support systems for NSCLC immunotherapy as both dedicated quantum algorithms evolve and physical hardware matures. Furthermore, we found supporting evidence that heavily reducing the feature space can improve generalizability without compromising performance. Future research is required to assess scalability to other clinical centers and validate these models on physical photonic quantum processors under realistic hardware noise conditions.

## 1 Introduction

Immune checkpoint inhibitors (ICIs) have transformed the management of non-small cell lung cancer (NSCLC), but the field still lacks robust biomarkers that can reliably predict which tumors will respond.

The only validated marker guiding ICI treatment in oncogene-wild type NSCLC is the tumor proportion score (TPS) of PD-L1 expression, yet this test is hampered by sampling limitations and substantial spatial and temporal heterogeneity across lesions and over time [Ilie et al., 2018, Hwang et al., 2021] with a modest overall discriminative performance with reported AUC values around 0.65 [Lu et al., 2019]. As a result, there is strong motivation to identify alternative or complementary biomarkers that can improve predictive performance while remaining clinically practical and minimally invasive [Genova et al., 2023].

Radiomics has emerged as a promising approach to address this need. By extracting high-dimensional quantitative features from standard imaging, radiomics can capture information about tissue architecture and tumor microenvironment that is often invisible to the human eye [Gillies et al., 2016]. In computed tomography (CT), these features reflect the interaction between X-rays and the biological properties of tissue, offering a noninvasive window into heterogeneity within and across lesions. Prior studies have demonstrated that radiomic signatures can predict pathological response, survival, and immunotherapy outcomes in NSCLC, indicating that imaging-derived features may serve as predictive biomarkers [Coroller et al., 2017, Wu et al., 2023, Trebeschi et al., 2019, Khorrami et al., 2020].

A key clinical need is the ability to predict treatment response at the level of individual lesions, a capability that could refine personalization for both advanced and early-stage NSCLC, and better align with contemporary strategies such as local consolidative radiotherapy in oligometastatic disease [Petrelli et al., 2018]. The present study builds on this literature while adopting a distinct focus: it aims to predict immunotherapy response on a per-lesion basis by combining CT radiomic features with novel photonic quantum machine learning architectures. This lesion-centric objective differs from much of the prior work, which primarily assessed patient-level outcomes or global response [Trebeschi et al., 2019, Khorrami et al., 2020, Prelaj et al., 2024]. The premise is that each lesion may reflect unique biological states, influenced by factors such as vascularization, immune infiltration, and stromal composition, leading to heterogeneous treatment responses within the same patient. Consequently, response in one lesion may not reliably indicate response across the full disease burden or overall survival. A lesion-level model, therefore, could provide more granular decision support, potentially guiding targeted interventions, adapting treatment strategies over time, and improving monitoring without the need for repeated invasive biopsies.

In summary, we argue that current biomarkers like PD-L1 TPS remain insufficient for precise response prediction in NSCLC, while radiomics offers a scalable, less invasive alternative that captures intra-patient heterogeneity. While standard classical machine learning is commonly applied to radiomic data, quantum machine learning (QML) offers a fundamentally different mathematical paradigm. By encoding classical data into quantum states, QML models map features into an exponentially large Hilbert space, providing a unique “inductive bias” that evaluates correlations and draws decision boundaries differently than classical networks. Therefore, this study proposes to evaluate whether an ideal, simulated photonic quantum machine learning model can predict immunotherapy response at the level of individual lesions, advancing beyond patient-level prognosis toward a more refined, lesion-specific framework. By establishing this theoretical baseline, we aim to demonstrate if this novel analytical approach can support more personalized immunotherapy strategies for NSCLC patients [Silvia Pamparino et al., 2026].

## 2 Methods

### 2.1 Research quality

The [METhodological RadiomICs Score](https://insightsimaging.springeropen.com/articles/10.1186/s13244-023-01572-w) (METRICS) of our study was 86.1% which corresponds to an “Excellent” Quality Category (Table S1).

### 2.2 Study Design and Setting

This retrospective multi-center study included adult patients with a histological diagnosis of advanced NSCLC who had been treated with an antiPD1 monotherapy as first or second line in three different public hospitals in Italy (Figure 1):

- Train Hospital: San Martino Polyclinic Hospital (Train Hospital, Genoa IT);
- Test Hospital 1: Parma Hospital (Test Hospital 1, Parma IT);
- Test Hospital 2: Messina Hospital (Test Hospital 2, Messina IT).

**Figure 1:**
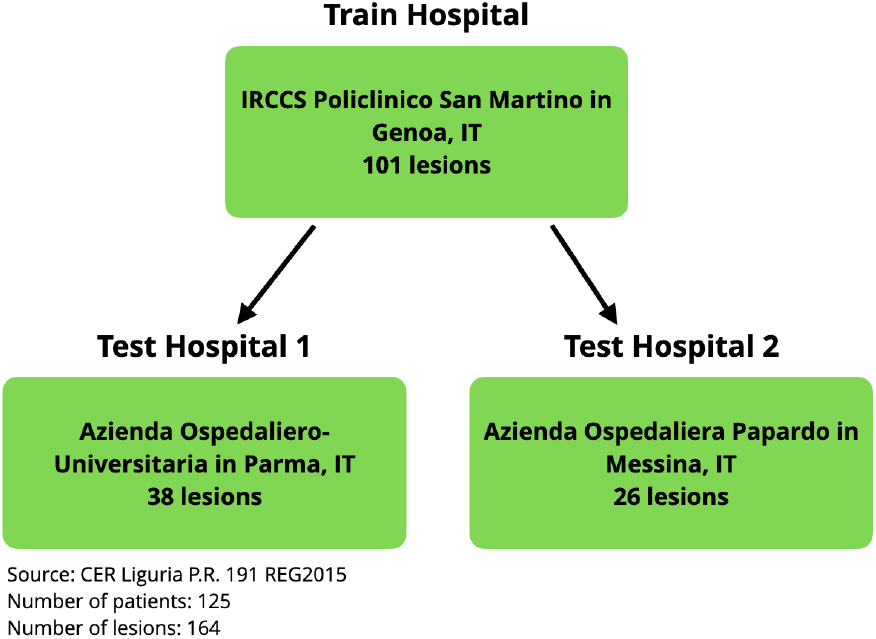
Study Design and Setting

### 2.3 Participants

The study included 125 patients (>= 18 years old) with 164 advanced NSCLC single lesions treated with anti-PD1 monotherapy as first or second line divided in three cohorts:

- train cohort of 101 lung lesions enrolled at the San Martino Polyclinic Hospital (Train Hospital), Genoa IT;
- test cohort 1 of 37 lung lesions enrolled at the Parma Hospital (Test Hospital 1), Parma IT;
- test cohort 2 of 26 lung lesions enrolled at the Messina Hospital (Test Hospital 2), Messina IT.

### 2.4 Procedures and Interventions

A baseline contrast-enhanced CT scan (chest-abdomen-pelvis protocol) performed within 30 days before starting anti-PD1 therapy was required. Each patient provided written informed consent for the collection of clinical data and the transmission of radiological images for study purposes. All patients underwent a baseline CT scan before the start of the treatment, the lesions were semi-automatically segmented and labeled as “progressive” if their diameter increased by more than 10% and as “non-progressive” if their diameter decreased by 10% or more over the following 6 months Figure 2.

**Figure 2:**
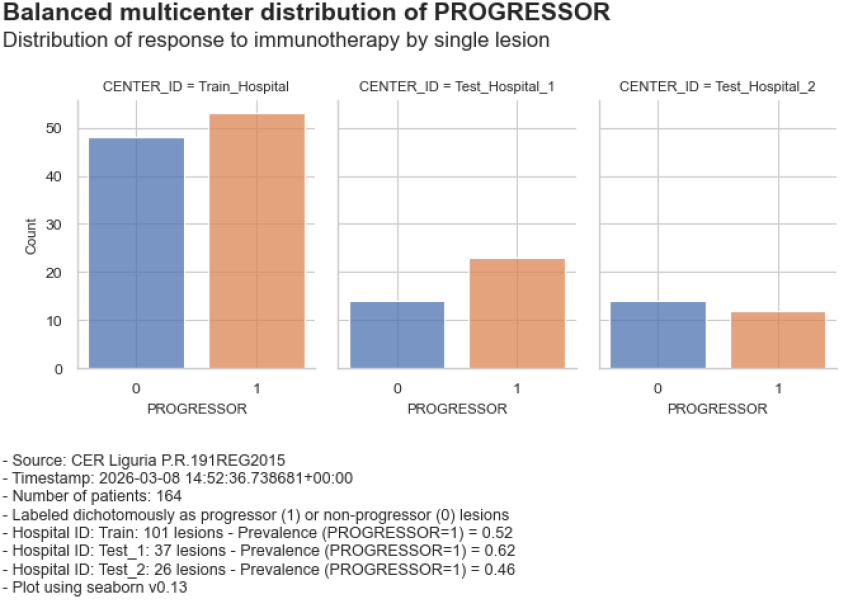
Distribution of target Progressor - Non Progressor

### 2.5 Segmentation and Radiomic Analysis

From each CT scan we extracted 851 radiomic features. Segmentation and features extraction were carried out using the same software in the different centers: segmentation was performed using 3D Slicer 4.10.1 and radiomic features were extracted with the Pyradiomics extension [Van Griethuysen et al., 2017]. The mathematical definitions of the adopted features are available for consultation on Pyradiomics.

### 2.6 Outcome Measures

1. The primary outcome measure of this study is the Average Precision Score of the radiomics-based photonic quantum machine learning models trained in the Train Hospital and tested in the Test Hospitals. Average Precision (AP) is a performance metric that summarizes a precision-recall curve into a single value, representing the average precision achieved at various recall levels. It calculates the weighted mean of precision scores at each threshold, where the weight is the increase in recall. AP measures model performance across all recall levels, with values ranging from 0 to 1 [Davis and Goadrich, 2006, Powers, 2011].
2. The secondary outcome measure is the number of evidence-based, robust and statistically significant features selected in the Train Hospital and utilized in the Test Hospitals. Feature selection is a preprocessing technique that identifies the key features of a given problem. It has traditionally been applied in a wide range of problems that include biological data processing and medical applications, where it can not only reduce dimensionality but also be a proxy metric for generalizability, i.e. a model using few selected and well understood radiomic features should generalize more easily than a model with hundreds of features [Remeseiro and Bolon-Canedo, 2019].

### 2.7 Statistical Analysis

In the Train Hospital:

- we identified 183 reliable features based on previous published clinical research;
- we selected the features with the highest robust skipped linear correlation and statistical significance at p<0.001;
- we trained 1 classical and 3 photonic quantum machine learning models.

In Test Hospitals 1 & 2, we tested the performance of the 4 models on unseen external datasets. Aiming to explore quantum machine learning as a long-term technology for precision oncology, we utilized an ideal classical simulation of the Open Access (OA) MerLin photonic quantum architecture. This approach assumes perfectly functioning hardware, eliminating confounding physical noise to assess true theoretical performance, while guaranteeing the perfect methodological reproducibility required in medical research.

#### 2.7.1 Why feature selection only in the Train Hospital?

Technology-rich domains like radiomics generate inherently noisy [Tu et al., 2021] high-dimensional data with few samples, which are valuable for biomarker discovery yet risk biased ML accuracy. Simulations show that standard K-fold CV remains overly optimistic even at n=1000 and small samples inflate reported performance. However, leakage from feature selection on pooled data contributes more to bias than hyperparameter tuning, and bias also depends on dimensionality, search space, and fold count [Vabalas et al., 2019]. For this reason we selected the most reliable and significant features in the Train Hospital and not on pooled data.

#### 2.7.2 Why skipped correlation

Previous research has analyzed various methods to reduce the feature space of high dimensional data for quantum computing. Among the various techniques:

- [Nembrini et al., 2021] and [Nau et al., 2025] approach feature selection as an optimization problem and solve it using a quantum annealing computer;
- [Felefly et al., 2023] developed an accurate interpretable quantum neural network model with quantum-informed feature selection;
- [Li et al., 2024] propose three graph-theoretic feature selection (GTFS) methods, including minimum cut (MinCut)-based, densest k-subgraph (DkS)-based, and maximal-independent set/minimal vertex cover (MIS/MVC)-based,

However, these approaches are performance-driven and based on machine learning algorithms, not evidence-driven and based on robust medical statistics.

On the other hand, skipped correlation is a robust statistical method used to measure linear association while ignoring outliers, serving as a robust alternative to Pearson’s correlation. It involves estimating the data’s center using the Minimum Covariance Determinant (MCD) and identifying outliers, often utilizing methods highlighted in National Institutes of Health (NIH) research [Wu and Kacker, 2021, Wilcox, 2003, Wilcox et al., 2018, Wilcox, 2004].

Unlike standard correlation, skipped correlation is less sensitive to extreme values, making it ideal for skewed data Figure 3.

**Figure 3:**
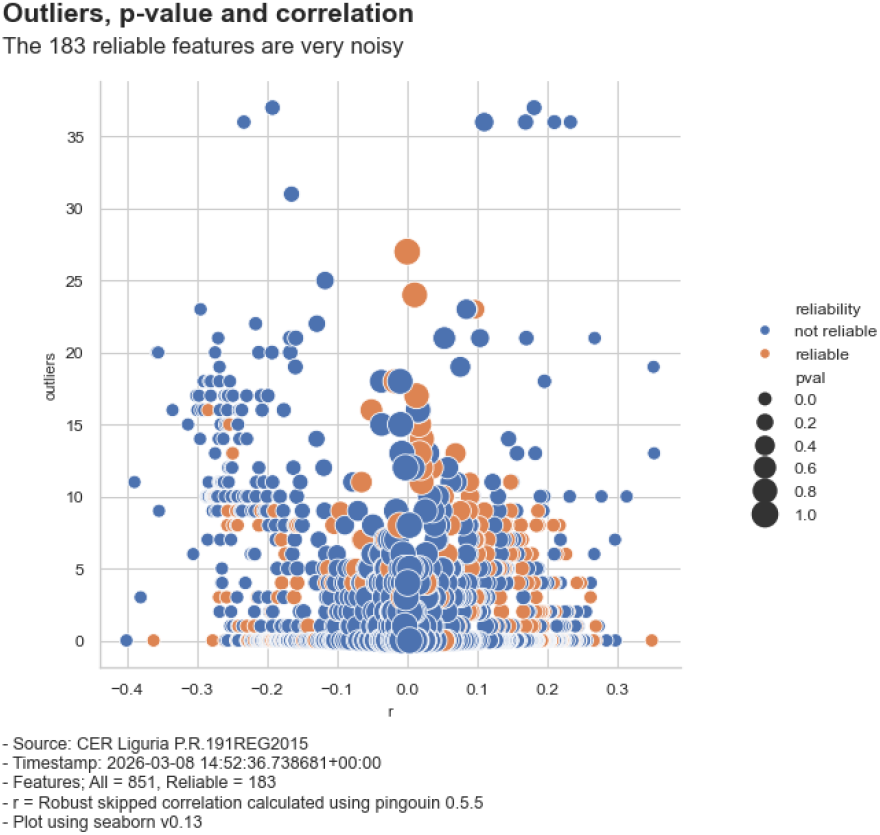
Distribution of the Outliers

### 2.8 Classical and Photonic Quantum Architectures

For the experimental implementation, we utilized the [MerLin: A Discovery Engine for Photonic and Hybrid Quantum Machine Learning](https://merlinquantum.ai/index.html) framework.

To ensure our experiment is fully transparent and reproducible, we adapted and significantly expanded the open-access [Quantum-Classical Hybrid Neural Network Comparison](https://merlinquantum.ai/notebooks/iris_classification.html) notebook. MerLin enables researchers to reproduce and build upon published quantum machine learning research. This section provides implementations of key papers like [Gan et al., 2022] in the quantum ML field, complete with working code, analysis, and extensions.

In particular, our reproduction includes:

- **Original code reproduction**: recreation of theMerLin’s methodology with modifications;
- **Reproduction Status**: the reproduction is complete with modifications;
- **Performance analysis**: we modified performance analytics to account for the clinical importance of recall.

More specifically, we implemented the following critical methodological modifications to transition from a general-purpose classification example to a robust clinical validation pipeline:

- **Data Pipeline Integration:** We replaced the standard Iris dataset with a specialized medical imaging pipeline, incorporating automated CSV ingestion, clinical target remapping (Progressor vs. Non-Progressor), and multi-center data partitioning across three distinct hospital cohorts;
- **Feature Reduction Workflow:** Unlike the reference code, we implemented an evidence-based feature-reduction stage utilizing robust medical statistics (skipped correlation) to filter features based on statistical significance (*p <* 0.001) and clinical reliability;
- **Architectural Adaptation:** We modified the core model construction to be dynamic, allowing for variable input and output dimensions. We specifically adapted the loss functions and output mapping strategies of both classical and quantum architectures to optimize for binary clinical targets;
- **Clinical Metric Expansion:** Evaluation was expanded beyond simple accuracy to include precision, recall, Average Precision (AP), and F1 scores, allowing for model selection criteria that account for the clinical implications of false positives;
- **Multi-Center Validation Flow:** We redesigned the execution logic from a single train-test split to a robust multi-center validation flow, where models are trained on a primary hospital and evaluated independently on two held-out external centers;
- **Reporting Standards:** We implemented clinical-grade visualization exports, including timestamped metadata, LaTeX table generation for neural network settings, and concatenated performance reporting for multi-run statistical stability.

As noted by the framework’s developers, practical gains in near-term quantum machine learning will be found through systematic, empirical evaluation across models, datasets, and hardware constraints [Notton et al., 2026].

All modifications in the MerLin code are the sole responsibility of the authors of this paper.

#### 2.8.1 Libraries

We utilized the following neural networks libraries for our experiment:

- PyTorch: For neural network implementation and training
- Perceval: For quantum circuit simulation
- Custom modules: For quantum layers and MLP models

#### 2.8.2 Photonic Quantum Architectures

We tested 4 model architectures to compare classical and quantum approaches:

1. Classical Models:
  a. MLP: Multi-layer perceptron with 8 hidden units, ReLU activation, and 0.1 dropout
2. Quantum Models:
  a. LINEAR-6modes: Uses linear output mapping with 6 quantum modes.
  b. LEXGROUPING-6modes: Uses lexicographic grouping strategy for output mapping
  c. MODGROUPING-6modes: Uses modular grouping strategy for output mapping

#### 2.8.3 Photonic Quantum Circuits

The quantum circuit is composed of three main sections:

1. Left interferometer (WL): Performs initial quantum state transformation using beam splitters and phase shifters
2. Variable phase shifters: Encodes the 2 input features into quantum phases. To bridge the dimensionality gap between the 2 selected radiomic features and the 6 spatial modes of the photonic circuit, we employed a zero-padding encoding strategy. The 2 continuous features were normalized and encoded as angles into the first two variable phase shifters. The remaining four phase shifters were padded with constant zero values, effectively embedding the classical medical data into the quantum state space without introducing artificial variance.
3. Right interferometer (WR): Performs final quantum state transformation

The interferometers use a rectangular arrangement of optical elements, where each layer consists of beam splitters (BS) and programmable phase shifters (PS). The phase shifters contain trainable parameters that are optimized during training via classical backpropagation. The depth of these interferometric layers and the total number of trainable phase shifts dictate the expressivity of the quantum model, acting analogously to the hidden weights in a classical neural network.

More information is available for consultation on MerLin: Quantum Expert Area.

#### 2.8.4 Training Parameters

We implemented a standard supervised learning loop with the characteristics described in Table 1.

**Table 1:** Parameters of the Neural Networks.

|  | Value |
| --- | --- |
| Input-size | 2 |
| Output-size | 2 |
| Optimizer: | Adam with lr 0.02 |
| Loss function | Cross-entropy |
| Batch size | 32 samples |
| Epochs | 50 (default) |
| Training samples | 101 |
| Features | 2 |
| Classes | 2 |
| Test 1 samples | 37 |
| Test 2 samples | 26 |

#### 2.8.5 Multi-Run Training Function

To ensure robust and statistically meaningful results, we trained each model variant multiple times with different random initializations. This approach helped us:

- Understand the variance in model performance
- Identify which architectures are more stable
- Avoid drawing conclusions from lucky/unlucky single runs
- Calculate confidence intervals for performance metrics

The function tracks both individual run results and aggregate statistics across all runs.

#### 2.8.6 Run the Complete Experiment

The complete experimental pipeline was executed as follows:

1. Multiple training runs: Each model variant is trained 5 times with different random seeds
2. Performance tracking: All metrics are recorded for statistical analysis
3. Visualization: Training curves and confusion matrices are generated
4. Statistical summary: Detailed performance statistics are computed

This comprehensive approach ensures reliable and reproducible results.

## 3 Results

Statistical analysis in the Train Hospital identified 2 features out of 851 that met the strict criteria for reliability and robust linear correlation with the target (*p <* 0.001). These features served as the input for both the classical and photonic quantum architectures.

As detailed in Table 3, all evaluated photonic quantum models exceeded the chance level (defined by local progressor prevalence) across both external validation cohorts. In Test Hospital 1, the LEXGROUPING-6modes model demonstrated superior performance compared to the optimized classical MLP baseline, achieving an Average Precision (AP) of 0.755 compared to 0.702. In Test Hospital 2, the same quantum architecture achieved performance parity with the classical baseline (AP 0.670).

These results, along with the performance of the LINEAR and MODGROUPING strategies, are summarized in Table 3. Detailed visualizations of the training stability, convergence speeds, and precision-recall metrics are provided in Appendix A (Training Curves Visualization) and Appendix B (Precision-Recall Curves Visualization).

### 3.1 Participant Characteristics

The prevalence of Progressor (label = 1) is shown in Table 2.

**Table 2:** Prevalence of Class Progressor (label = 1)

|  | Lesions | Progressor<br>Prevalence |
| --- | --- | --- |
| Train Hospital | 101 | 0.52 |
| Test Hospital 1 | 37 | 0.62 |
| Test Hospital 2 | 26 | 0.46 |

### 3.2 Primary Outcomes

The classical and photonic quantum architectures achieved the performances in Test Hospitals 1 & 2 described in Table 3.

**Table 3:** Results.

| Center | Model Name | Progressor Prevalence | Average Precision Score |
| --- | --- | --- | --- |
| Test Hospital 1 | Classical Multi-Layer Perceptron (MLP) | 0.622 | 0.702 |
| Test Hospital 1 | LINEAR-6modes-nobunching | 0.622 | 0.697 |
| Test Hospital 1 | LEXGROUPING-6modes | 0.622 | 0.755 |
| Test Hospital 1 | MODGROUPING-6modes | 0.622 | 0.655 |
| Test Hospital 2 | Classical Multi-Layer Perceptron (MLP) | 0.462 | 0.670 |
| Test Hospital 2 | LINEAR-6modes-nobunching | 0.462 | 0.657 |
| Test Hospital 2 | LEXGROUPING-6modes | 0.462 | 0.670 |
| Test Hospital 2 | MODGROUPING-6modes | 0.462 | 0.661 |

### 3.3 Secondary Outcomes

Table 4 shows that in Train Hospital we identified 2 features out of 851 that were robustly correlated to the target with statistical significance at p<0.001.

**Table 4:** All features with a pvalue < 0.001.

| features | outliers | r | pval | power | reliability |
| --- | --- | --- | --- | --- | --- |
| ('wavelet-LLH', 'firstorder', 'TotalEnergy') | 7 | -0.40 | 0.00 | 0.98 | not reliable |
| ('wavelet-HLL', 'ngtdm', 'Busyness') | 12 | -0.40 | 0.00 | 0.98 | not reliable |
| ('wavelet-HHH', 'glrlm', 'LongRunLowGrayLevelEmphasis') | 11 | -0.39 | 0.00 | 0.97 | not reliable |
| ('wavelet-LLL', 'glcm', 'Imc1') | 0 | -0.36 | 0.00 | 0.96 | reliable |
| ('wavelet-LLH', 'firstorder', 'Energy') | 7 | -0.37 | 0.00 | 0.96 | not reliable |
| ('wavelet-LLL', 'glcm', 'Imc2') | 0 | 0.35 | 0.00 | 0.95 | reliable |
| ('wavelet-HHL', 'glrlm', 'ShortRunLowGrayLevelEmphasis') | 20 | 0.37 | 0.00 | 0.93 | not reliable |
| ('wavelet-HHH', 'glrlm', 'ShortRunLowGrayLevelEmphasis') | 13 | 0.35 | 0.00 | 0.93 | not reliable |

#### 3.3.1 Feature Space Reduction

Table 5 shows that only a subgroup of 2 radiomic features were both reliable and linearly robustly correlated to the target with statistical significance at p<0.001.

**Table 5:** Reliable features with a pvalue < 0.001.

| features | outliers | corr | p-value | power | reliability |
| --- | --- | --- | --- | --- | --- |
| ('wavelet-LLL', 'glcm', 'Imc1') | 0 | -0.36 | 0.00 | 0.96 | reliable |
| ('wavelet-LLL', 'glcm', 'Imc2') | 0 | 0.35 | 0.00 | 0.95 | reliable |

See also Figure 4 and Figure 5

**Figure 4:**
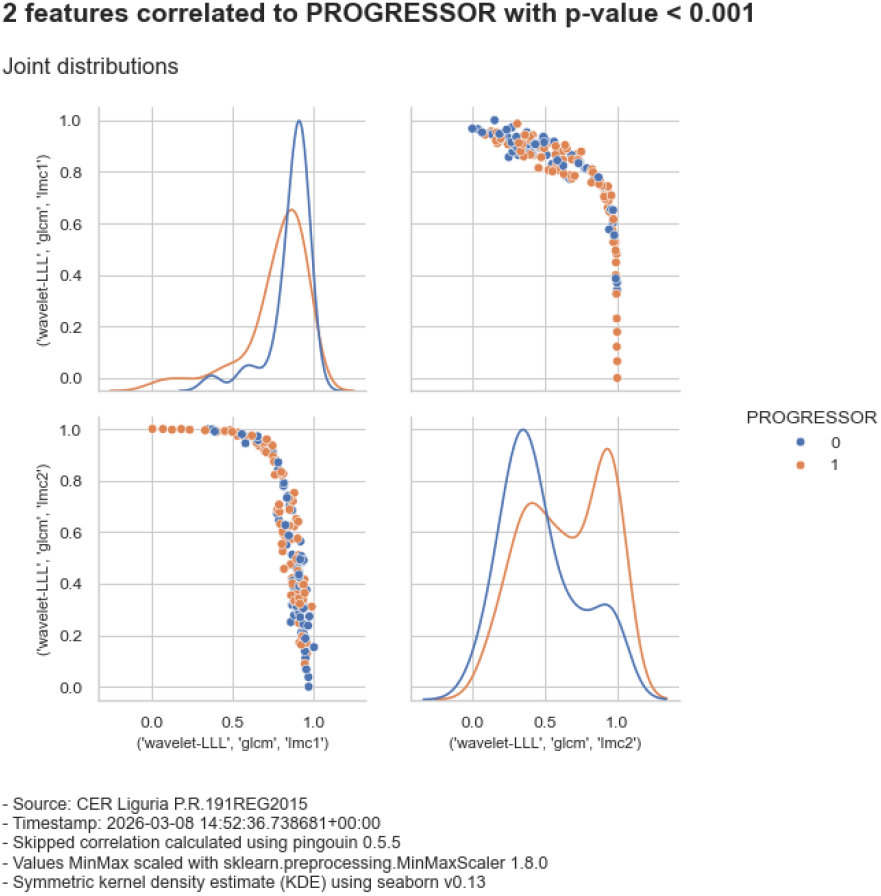
Distribution of the 2 reliable and statistically significant features

**Figure 5:**
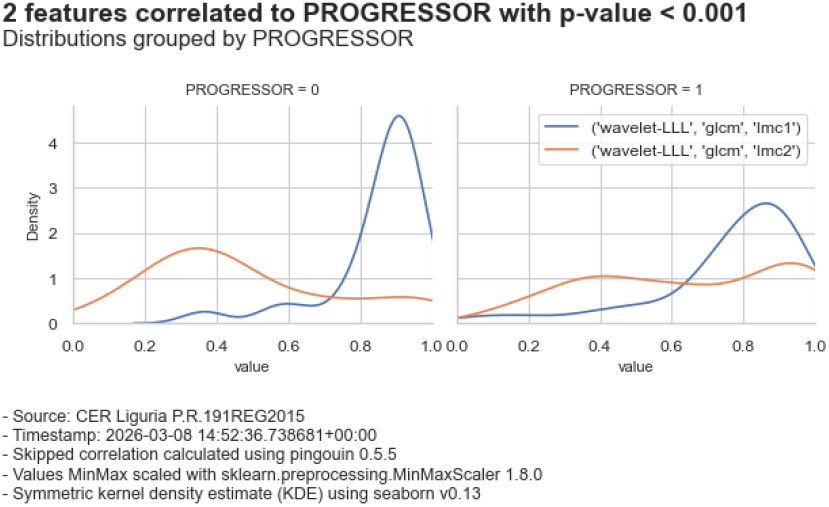
Distribution of the 2 reliable and statistically significant features by Progressor

## 4 Innovation

To our knowledge, this is the first study to test the external validity of radiomics-based photonic quantum architectures utilizing an evidence-based, statistically significant reduced feature space. By demonstrating that an ideal, simulated quantum model (LEXGROUPING-6modes) can explicitly outperform or perfectly match a highly optimized classical neural network across unseen multi-center datasets, this work establishes a critical theoretical benchmark. It proves that quantum architectures possess the native mathematical expressivity to capture complex biological signals relevant to NSCLC immunotherapy response.

Methodologically, the innovation of this study is twofold. First, we provide empirical evidence that aggressively reducing a high-dimensional radiomic space (from 851 down to 2 strictly reliable features) via robust medical statistics not only prevents overfitting but actively enables superior generalizability. Second, by utilizing an open-access simulated framework (MerLin) on classical hardware, we ensure the perfect methodological reproducibility of these findings, effectively isolating the models’ true predictive power from the confounding limitations of today’s noisy quantum hardware.

These findings strongly support the long-term viability of quantum machine learning as a precision medicine tool. Future research must assess scalability to larger clinical cohorts and validate these specific architectures on physical photonic quantum processors under realistic hardware noise conditions,specifically accounting for photon loss, detector dark counts, and imperfect photon indistinguishability, as both dedicated quantum algorithms and physical hardware continue to mature.

## 5 Discussion

### 5.1 Principal Findings

Based on the experimental results, we can draw several key conclusions:

- radiomic features, when evaluated at the lesion level, contain robust and actionable biological signals for predicting immunotherapy response in advanced NSCLC;
- the inherently high-dimensional radiomics feature space can be aggressively and effectively reduced (from 851 to 2 features) using strict, evidence-based statistical thresholds (*p <* 0.001), which actively prevents overfitting and preserves multi-center generalizability;
- simulated photonic quantum machine learning models not only compete with but can explicitly outperform optimized classical neural networks on unseen external datasets. Demonstrating this capability is a significant milestone, confirming that QML holds genuine promise as a precision medicine tool as both algorithms and physical hardware mature.

Notably, the superior performance of the LEXGROUPING-6modes architecture provides a compelling mathematical insight into quantum-classical data mapping. While modular grouping (parity-based) and classical linear mappings showed competitive baseline performance, the lexicographic binning strategy explicitly outperformed the classical MLP in Test Hospital 1 (Average Precision 0.755 vs. 0.702) and perfectly matched it in Test Hospital 2 (0.670).

This suggests that a dictionary-based, hierarchical sorting of the quantum output states naturally aligns with the complex decision boundaries of the encoded radiomic features. By preserving the structured distribution of the photon measurements—rather than smoothing or averaging them through classical weights—the lexicographic strategy effectively leveraged the quantum model’s unique inductive bias to capture true biological signals.

### 5.2 Comparison with Existing Literature

The models show a tendency to favor the progressor class: this produces high recall but lower precision, increasing false positives. Methodologically, this is a weakness, but clinically it may be an advantage because it prioritizes early identification of lesions at risk of progression, enabling prompt local treatments such as stereotactic radiotherapy.

The study emphasizes that the models are not intended to replace clinical judgment or to predict patient survival; instead, they provide lesion-specific predictions that can inform multidisciplinary decision-making. This lesion-level perspective aligns with how clinicians manage heterogeneous disease, where selective local treatment of resistant lesions is feasible while systemic immunotherapy continues.

### 5.3 Strengths and Limitations

We are candid about the limitations of this study. The cohort size is relatively small, which reduces statistical power and limits the diversity of phenotypes seen during training. Segmentation is semi-automatic or manual and performed by multiple radiologists, introducing variability that can affect feature extraction and accuracy. This variability may also improve generalizability by exposing the model to realistic inter-reader differences.

The lesion-level strategy introduces statistical dependence because multiple lesions can come from the same patient, sharing biological and imaging characteristics. This dependence complicates evaluation and can inflate performance if not accounted for statistically.

The model also misclassifies approximately 30 percent of responses, which limits immediate clinical deployment. However, this performance is comparable to PD-L1 tissue expression testing, with the advantage that radiomics does not require tissue samples from each lesion and can be applied broadly.

Intrinsic limitations of radiomics are highlighted. Accurate segmentation is essential, yet some lesions are difficult to segment due to imaging artifacts, atelectasis, or proximity to the pulmonary hila. This observation underscores the importance of segmentation quality and suggests that lesion type and anatomical context influence radiomic reliability. It also implies that future pipelines should quantify contour uncertainty and consider lesion-specific stratification to improve performance and interpretability.

Furthermore, CT-based features operate at a voxel scale and cannot capture cellular-level or microenvironmental biology directly, constraining biological specificity. These limitations set a performance ceiling and motivate the need for complementary information sources. We propose larger and more heterogeneous training cohorts to improve generalizability, along with automated segmentation pipelines to reduce variability.

We also suggest integrating clinical covariates such as PD-L1 expression, neutrophil-to-lymphocyte ratio (NLR), platelet-to-lymphocyte ratio (PLR), demographics, smoking status, and BMI to add orthogonal predictive signals. Circulating biomarkers such as extracellular vesicle (EV) PD-L1 and circulating tumor DNA (ctDNA) dynamics are highlighted as promising additions that could improve predictive accuracy and reduce false positives. These enhancements would shift the model from a purely imaging-based predictor to a multimodal decision-support system.

Future evaluations should include calibration analyses, transparent reporting of subgroup performance, and prospective validation with predefined endpoints. We emphasize that lesion-level predictions must be integrated into clinical workflows thoughtfully, with clear decision thresholds and an understanding of how predictions affect resource allocation and treatment planning. Prospective studies should assess whether model-guided local therapy improves outcomes or simply shifts clinical workload. Governance around how discordant lesion predictions are adjudicated will also be necessary for safe deployment.

Last, the SPIRIT-AI and CONSORTAI initiatives provide guidance to improve the transparency and completeness of reporting of clinical trials evaluating interventions involving artificial intelligence [Ibrahim et al., 2021, Martindale et al., 2024, Liu et al., 2020, Rivera et al., 2020]. The utility of these initiatives is not clear for use in Quantum Machine Learning and future research is required to assess scalability to other clinical centers and validate these models on physical photonic quantum processors under realistic hardware noise conditions.

### 5.4 Clinical and Research Implications

This study evaluates the real-world applicability of radiomics for predicting immunotherapy response in patients with advanced non-small cell lung cancer (NSCLC). Four predictive models are developed and are tested on two external validation datasets. A central design decision is to analyze disease at the lesion level rather than the patient level, motivated by intra-patient heterogeneity in response patterns. This framing mirrors clinical practice, where individual lesions may behave differently and may require distinct local interventions while systemic therapy continues. Although performance declines modestly from internal to external evaluation, the consistency across sites suggests that the models generalize across institutions, imaging protocols, segmentation workflows, and reader variability. [Trebeschi et al., 2019, Khorrami et al., 2020, Wu et al., 2023, Coroller et al., 2017, Prelaj et al., 2024]

A notable aspect is the exclusive use of radiomic features without clinical variables. This constraint is acknowledged as a limitation, yet it also tests whether imaging alone can provide meaningful predictive information. Radiomic features can be extracted noninvasively from standard CT scans, making the approach scalable and applicable to every lesion without repeated biopsies [Gillies et al., 2016].

## 6 Conclusion

In conclusion, this study demonstrates that combining lesion-level radiomic analysis with photonic quantum machine learning offers a highly promising, non-invasive approach for predicting immunotherapy response in advanced NSCLC. Crucially, by rigorously reducing the high-dimensional feature space to just two robust biological signals, our simulated quantum architecture (LEXGROUPING-6modes) explicitly outperformed or perfectly matched an optimized classical neural network across unseen, multi-center external validation datasets.

This lesion-centric framework aligns seamlessly with contemporary clinical practice. By capturing intra-patient response heterogeneity, it provides the granular decision support necessary to enable selective local treatments (e.g., radiotherapy) for non-responding lesions while systemic therapy continues. As a scalable and perfectly reproducible layer of evidence, these quantum-assisted radiomic models can complement existing biomarkers like PD-L1, supporting longitudinal monitoring without the need for repeated invasive biopsies.

Despite inherent limitations related to sample size, segmentation variability, and the deliberate use of an ideal noiseless simulation, this work establishes a rigorous theoretical baseline. It outlines a clear roadmap for the future: realizing the full clinical potential of this approach will require larger prospective cohorts, automated segmentation pipelines, and, critically, the transition from simulated environments to physical photonic quantum processors under realistic hardware noise conditions. With continued algorithmic evolution and hardware maturation, quantum-enhanced radiomics could become a transformative tool for personalized immunotherapy management.

## Author Declarations

All relevant ethical guidelines have been followed, and any necessary IRB and/or ethics committee approvals have been obtained.

## IRB/oversight

The details of the IRB/oversight body that provided approval or exemption for the research described are given below: This study was carried out in compliance with the rules of the Helsinki Declaration and International Ethical Regulations [Association et al., 2009], including all subsequent amendments, under the approval of the Research Ethics Committee of the IRCCS Azienda Ospedaliera Universitaria - Istituto Nazionale per la Ricerca sul Cancro, Genova - N. Registro CER Liguria: P.R. 191REG2015. All necessary patient/participant consent has been obtained and the appropriate institutional forms have been archived, and that any patient/participant/sample identifiers included were not known to anyone (e.g., hospital staff, patients or participants themselves) outside the research group so cannot be used to identify individuals. All clinical trials and any other prospective interventional studies must be registered with an ICMJE-approved registry, such as ClinicalTrials.gov. We confirm that any such study reported in the manuscript has been registered and the trial registration ID is provided (note: if posting a prospective study registered retrospectively, please provide a statement in the trial ID field explaining why the study was not registered in advance). All appropriate research reporting guidelines and uploaded the relevant EQUATOR Network research reporting checklist(s) and other pertinent material as supplementary files, if applicable, have been followed.

## Acknowledgements

We gratefully acknowledge Silvia Pamparino, Alessandro Fedeli, Giulia Mazzaschi, Alessandro Russo, Simone Caprioli, Enrica Teresa Tanda, Chiara Dellepiane, Martina Fiannacca, Edoardo Nizzi, Francesco Buemi, Gianluca Milanese, Maurizio Balbi, Lodovica Zullo, Rosanna Turrisi, Giuseppe Cittadini, Alberto Tagliafico, Nicola Sverzellati, Marcello Tiseo, and Marco Tagliamento for their valuable contributions to this research.

Furthermore, we acknowledge the MerLin framework utilized in this study. MerLin is a discovery research project supported by MITACS Accelerate (projects IT45761 and IT36314), the UFOQO Project financed by the French State as part of France 2030, the European Union’s Horizon Europe research and innovation programme under grant agreement No 101130384 (QUONDENSATE), and the QuantERA programme through the project ResourceQ.

Certain materials and code adaptations used in this manuscript are © of MerLin and/or Quandela, and their use is granted under the MIT License.

## Author Contributions

Conceptualization: SO, FS, EB; Methodology: SO, FS, EB; Software: SO, FS, DM; Validation:SO, FS, EB, GR, FG; Formal Analysis:SO, FS, EB; Investigation:SO, FS, EB, GR, FG, CG; Resources:FS, EB, DM; Data Curation: EB, GR, CG, FG; Writing - Original Draft: SO, FS; Writing - Review & Editing: SO, FS, CG, EB, GR, NH; Visualization: SO, FS, DM; Supervision:EB, FS, FG, CG; Project Administration: SO, FS, EB, CG; Funding Acquisition: FG, CG

## Data Availability

Data are available from the corresponding author upon reasonable request.

## A Training Curves Visualization

This section provides a comprehensive view of the training process by plotting four key metrics:

- Training Loss: Shows how well the model fits the training data
- Training Accuracy: Indicates the model’s performance on training samples
- Test Accuracy: Reveals the model’s generalization capability
- Test F1: Highlights the balance between precision and recall on the unseen external validation dataset

For each model variant, we display:

- Solid line: Mean performance across all runs
- Shaded area: Performance envelope (min to max values)

This visualization helps identify overfitting, convergence speed, and stability. The following plots show how each model’s performance evolves during training, specifically highlighting:

- Convergence speed: How quickly each model reaches its best performance
- Stability: Width of the shaded area indicates variance across runs
- Final performance: Where each model plateaus

**Figure 6:**
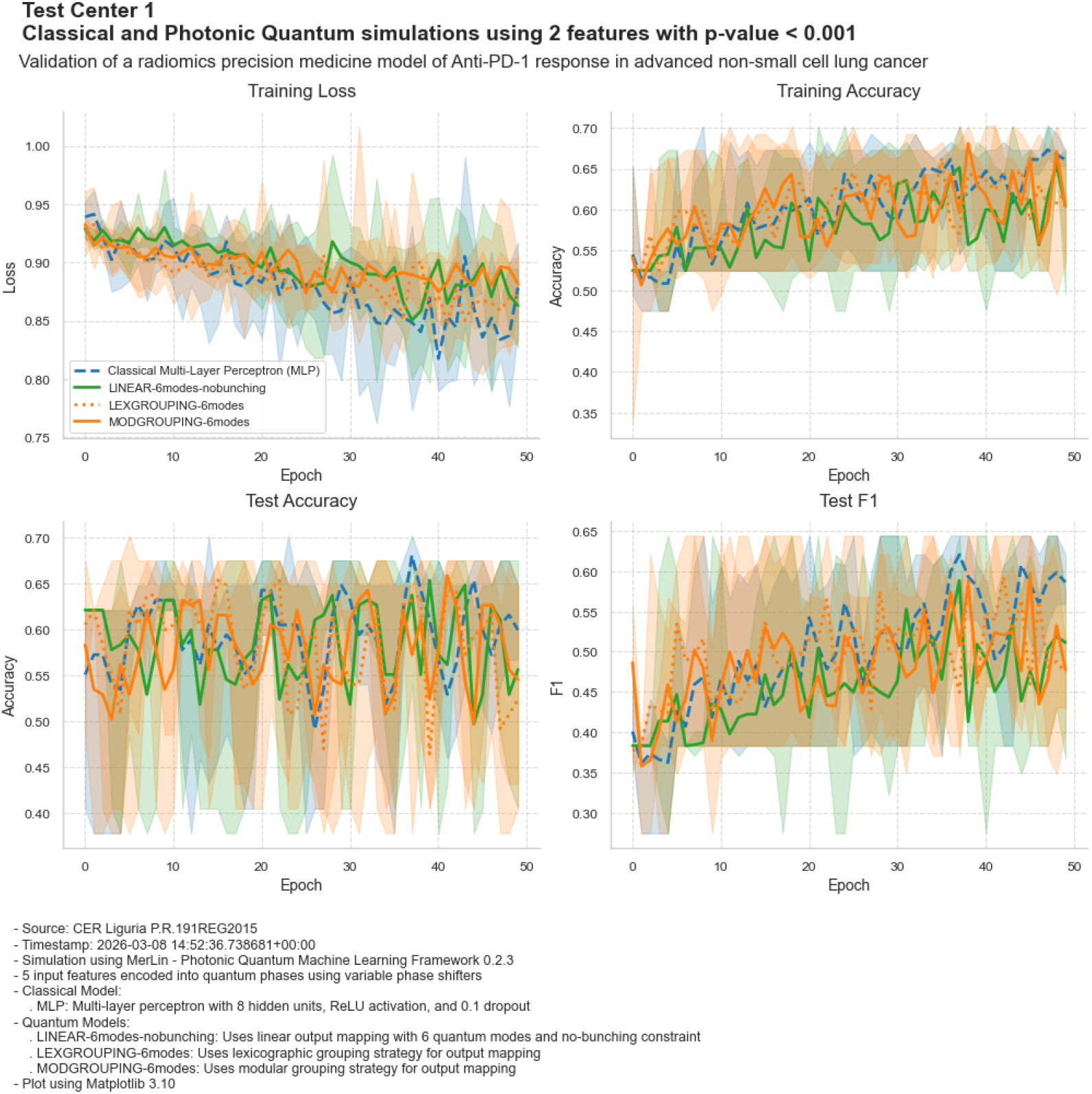
A comprehensive view of the training process in Train Hospital and external validation process in Test Center 1, plotting Loss, Accuracy, and F1 metrics.

**Figure 7:**
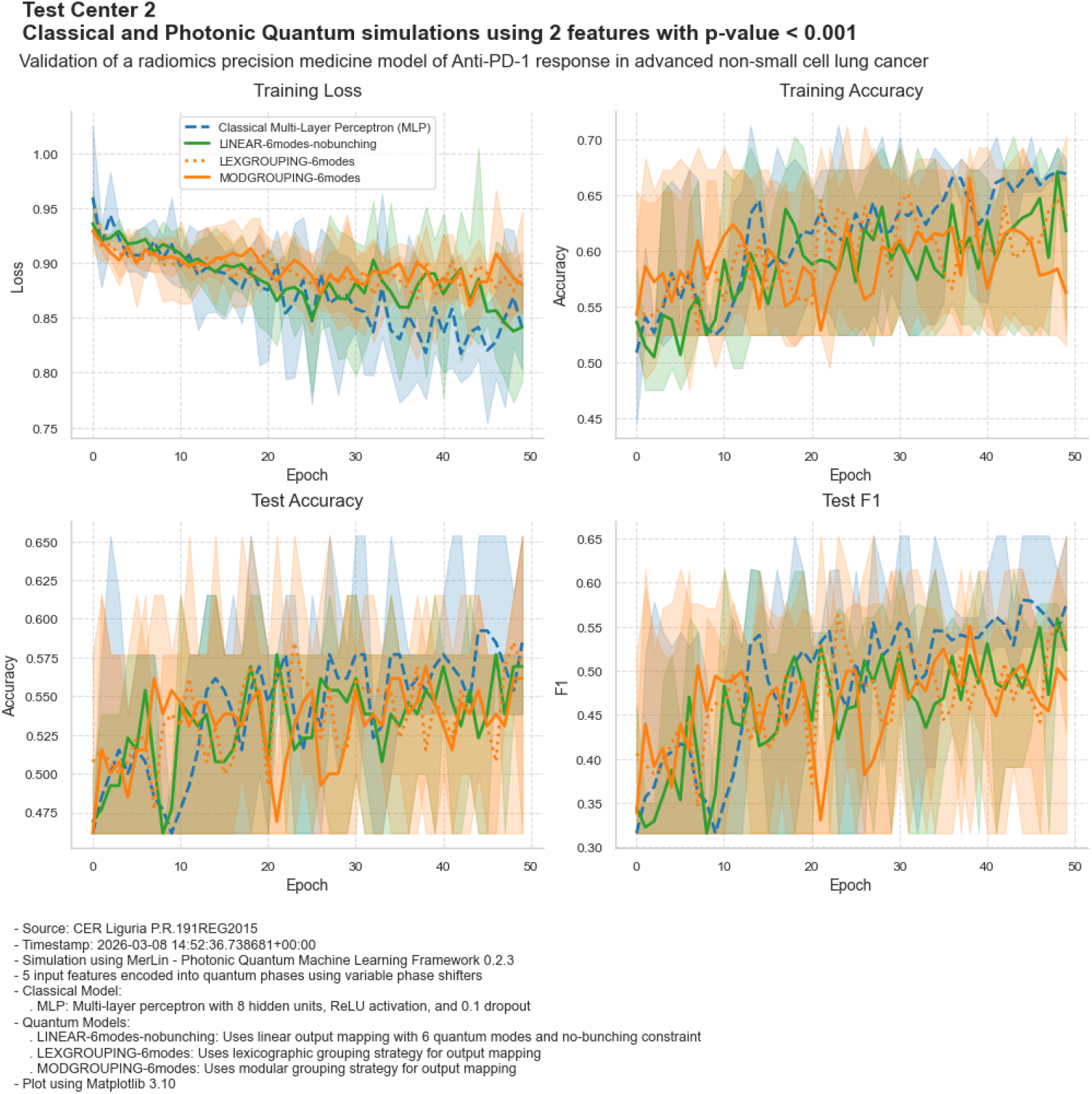
A comprehensive view of the training process in Train Hospital and external validation process in Test Center 2, plotting Loss, Accuracy, and F1 metrics.

## B Confusion Matrices and Precision-Recall Curves Visualization

The following visualizations detail the classification performance of the classical and quantum architectures on the external validation datasets. They present the confusion matrices alongside the precision-recall curves, Average Precision (AP) scores, and the chance level (progressor prevalence) for each evaluated model to provide a transparent view of false positives, false negatives, and overall model precision.

**Figure 8:**
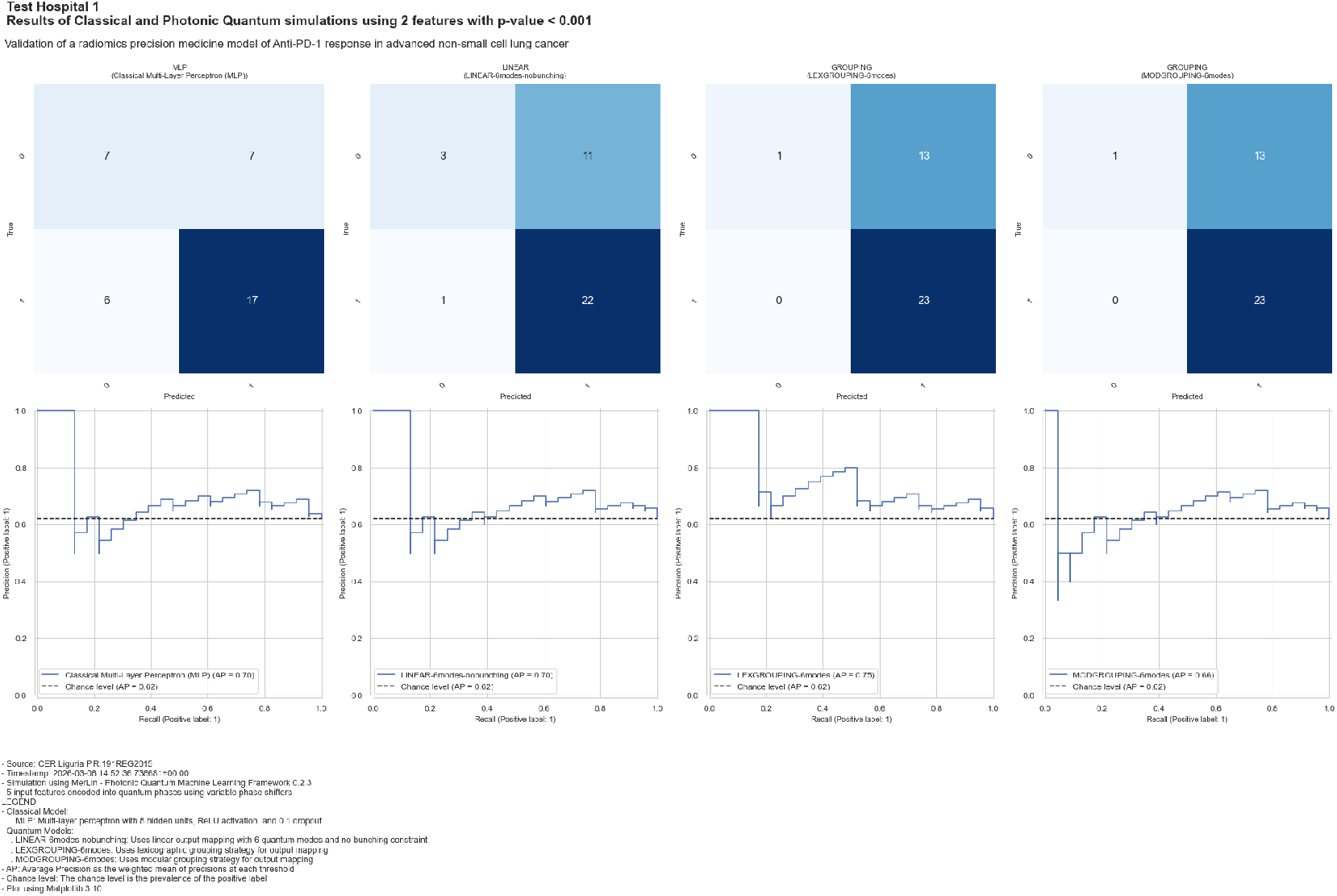
Confusion Matrices, Precision-Recall Curves, Average Precision Scores, and Chance Level (Progressor Prevalence) for Test Center 1

### Test Center 2

**Figure 9:**
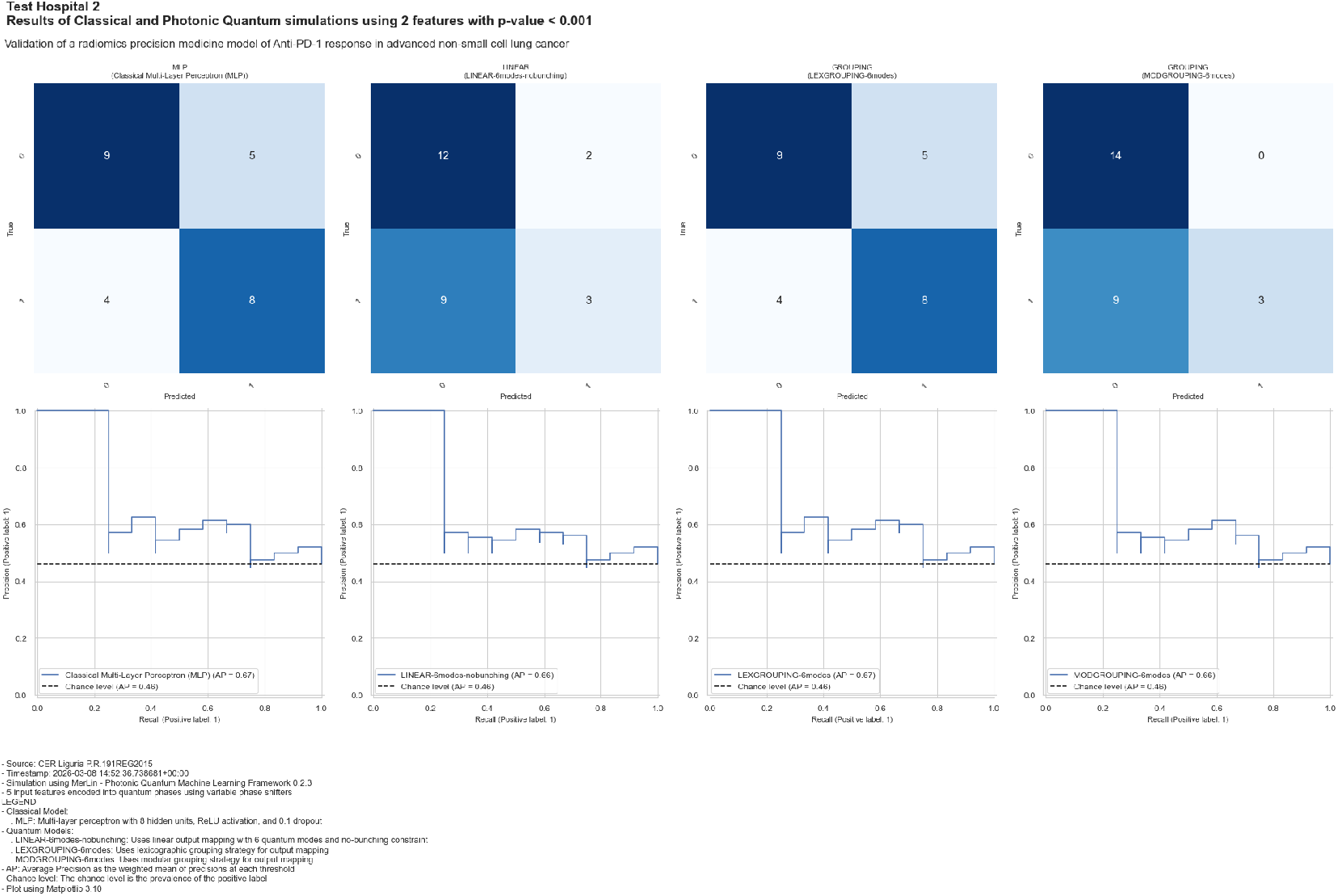
Confusion Matrices, Precision-Recall Curves, Average Precision Scores, and Chance Level (Progressor Prevalence) for Test Center 2

